# Normative data for the 10-min Lean Test in individuals without Orthostatic Intolerance

**DOI:** 10.1101/2025.05.06.25327047

**Authors:** Nafi Iftekhar, Amy Wilson, Louis Nguty, Hussain Al-Hilali, Yusur Al-Hilali, Kinshuk Jain, Angela Braka, Thomas Osborne, Manoj Sivan

## Abstract

**Background:** Orthostatic intolerance syndromes such as Orthostatic Hypotension (OH) and Postural Orthostatic Tachycardia Syndrome (PoTS) are common symptoms seen in post-infection conditions and other neurological conditions with autonomic dysfunction. The 10-minute National Aeronautics and Space Administration Lean Test (NLT) is an objective clinical test used to assess these symptoms and direct management. There is, however, no robust literature on normative data for this test, particularly from a younger population.

**Aims:** The aim of this study was to produce a healthy control data set for NLT, which can be used for comparison with the patient population with health conditions.

**Methods:** Individuals recruited into the study had no history or symptoms of orthostatic intolerance; autonomic dysfunction; post-infection conditions (such as long COVID); or other neurological conditions with hemodynamic instability. Participants were primarily recruited from the general population in a metropolitan city. All participants underwent a standardised NLT. Lying Blood Pressure (BP) and Heart Rate (HR) after 2 min of lying down supine was recorded, followed by BP and HR recordings at every minute of standing (leaning against a wall) up to 10 minutes, along with recording patient-reported symptoms at each time point.

**Results:** A complete dataset was available for 112 individuals (60.7% Female, 39.3% Male). The population was 61.6% Caucasian, 8.0% Asian, 3.6% Black/Caribbean, 9.8% Mixed, and 17.0% Other; the mean age was 35.3 ± 15.1, with a BMI of 24.8 ± 4.0; 30.6% of individuals had a background medical condition, but none of the exclusion criteria.

During NLT, upon standing, the average change of HR was an increase of 9.89 ± 8.15bpm. The sustained HR increase (HR increase sustained at two consecutive readings) was an average of 6.23 ± 6.94bpm. The predominant response with BP was an increase of systolic BP, with the average initial increase being 7.55 ± 10.88mmHg. None of the participants met the diagnostic criteria for symptomatic OH or PoTS during NLT.

**Conclusion:** For the first time in the current literature, NLT data from a relatively younger healthy population without orthostatic intolerance have been gathered. This normative data will help interpret NLT findings in younger patients with Orthostatic Intolerance better and be useful in managing dysautonomia in specific conditions.

## Introduction

Orthostatic Intolerance (OI)(1) refers to symptoms arising from the inability to maintain normal blood pressure or heart rate when standing upright, which is then alleviated by reclining or lying down (2). OI can cause symptoms such as myalgia (3), dizziness, syncope (4), fatigue, headache, nausea, and palpitations (5, 6).

Postural orthostatic tachycardia syndrome (PoTS) (7) is a type of orthostatic intolerance characterised by an excessive increase in heart rate (HR) (more than 30 beats per minute or over 120 beats per minute) within the first 10 minutes of standing. Orthostatic Hypotension (OH) is characterised by a drop in systolic BP by more than 20 mmHg (or drop in diastolic BP by more than 10 mmHg) within the first 3 minutes of standing. If OH is present, a diagnosis of PoTS cannot be made.

Both PoTS and OH has been noted in literature to be associated with Long COVID (LC); a condition characterised by persistent symptoms, lasting greater than 12 weeks, experienced after recovering from an acute COVID-19 infection (8, 9). The common symptoms of LC are post-exertional malaise, fatigue, brain fog, pain and dizziness (10). In the UK, nearly 2 million individuals are reported with LC (11). In an average-sized medical practice in the UK, a general practitioner can anticipate having approximately 65 patients affected with LC (12, 13).

One of the plausible mechanisms in LC is dysautonomia; a term that refers to the dysfunction of the autonomic nervous system (ANS) (14, 15), which regulates involuntary bodily functions such as heart rate, blood pressure, digestion, sweating and temperature. Dysautonomia can have various causes, such as infections, autoimmune diseases, genetic disorders, or trauma (16). PoTS and OH are the common dysautonomia syndromes of the cardiovascular system (17-19).

There are a few proposed theories on pathophysiology of dysautonomia in LC, including direct damage to autonomic nervous system by the virus and indirect damage via autoantibodies (20). There is considerable recent evidence reporting a high prevalence of dysautonomia syndromes in LC (21). The prevalence for PoTS in LC is reported to be approximately 40-50% (22-24) and approximately 40% (24-26) of LC patients have been reported to have OH.

Dysautonomia is also observed in a range of medical conditions such as amyloidosis and HIV to Guillain– Barre Syndrome and paraneoplastic syndromes (27, 28). The OI syndromes of PoTS and OH have been reported in conditions such as Parkinson’s, Multiple Sclerosis and Diabetes mellitus (29, 30).

When assessing patients for OI syndromes, a range of objective tests can be used. The conventional test is the Head Up Tilt (HUT) table test which is carried out in hospital settings. A simpler clinic-based test is the National Aeronautics and Space Administration Lean Test (NLT) (13, 31), that can be conducted in a clinic or at home setting. Recent studies have validated the use of NLT for detecting OI syndromes in LC (32, 33). The need for normative data to interpret the findings of NLT has been highlighted in these studies.

The purpose of this study was to produce a normative data set from healthy volunteers, and from a younger population. This will enable us to interpret the NLT findings in medical conditions with greater confidence and manage the symptoms more comprehensively.

## Methods

### The LOCOMOTION Study

The work reported here was part of LOCOMOTION (LOng COvid Multidisciplinary consortium Optimising Treatments and services across the NHS), a 30-month multi-site case study of ten LC clinics beginning in 2021, which sought to optimise LC care across the clinics. The study protocol with details of management, governance and patient involvement has been previously published (34). Ethical approval was granted by Yorkshire & The Humber— Bradford Leeds Research Ethics Committee (REC; ref: 21/YH/0276) and subsequent amendments.

#### Patient sampling and consenting

The inclusion criteria for this study were individuals without a formal diagnosis of LC or any condition with dysautonomia or hemodynamic instability. Exclusion criteria for the study was the inability to give informed consent or comply with test instructions, if the clinical team considered the test unsuitable or unsafe (e.g., if the patient could not stand unaided), and any coexisting condition that could interfere with autonomic or hemodynamic function. Verbal and written informed consent to perform the NLT were obtained from all participants.

Patients were recruited in a 5-month period from a metropolitan city.

#### Sampling

The sampling frame for this study was purposive to match the demographics of LC patients seen in NHS clinics.

#### The NASA Lean Test

The NLT was performed according to published instructions (31, 33).

All participants first lay quietly for two to five minutes; one supine reading of HR and BP was obtained, which is used as a participant’s baseline. Subjects then stood slowly, shoulders leaning against a wall and feet positioned 15 cm from the wall. HR and BP measurements were obtained at standing (0 min) and then at one-minute intervals until 10 minutes of standing had been completed.

The NLT was terminated prematurely if participants’ symptoms were such that they were unable to complete the full 10 minutes or if the researcher was concerned for participant safety. If the NLT was stopped early, the test was still valid, and the available data were still analysed and standard criteria applied.

Below are the definitions for the possible results of the NLT:

- Orthostatic Intolerance (OI) symptoms: Dizziness/light-headedness, palpitations, chest pain, or tremulousness which presents or gets worse when sitting or standing and improves or resolves when lying down.
- Orthostatic Hypotension (OH): Fall in systolic blood pressure within the first three minutes of standing of at least 20 mmHg or fall in diastolic blood pressure of at least 10 mmHg with or without acute symptoms during the test.
- Late Orthostatic Hypotension (OH): Fall in systolic blood pressure after the first three minutes of standing of at least 20 mmHg or fall in diastolic blood pressure of at least 10 mmHg with or without acute symptoms during the test.
- Symptomatic Orthostatic Hypotension (OH): OH plus OI symptoms during the NLT
- Orthostatic Tachycardia (OT): Increase in HR above baseline of at least 30 bpm at any point during the 10 min or any HR reading equal or above 120 bpm.
- Postural Orthostatic Tachycardia Syndrome (PoTS): OT plus OI symptoms during the NLT
- Sustained OT/PoTS: Increase in HR that is maintained for at least two consecutive readings 1 min apart during the NLT.
- Symptomatic NLT: Any symptoms reported during the NLT, including baseline symptoms that were exacerbated by standing.
- Positive NLT: Patients meeting the criteria for POTS or symptomatic OH

#### Data Collection and Management

HR and BP were collected with the Omron M2 blood pressure monitor, which has been endorsed by the British Hypertension Society (35).

Data was aggregated through a form created on Jisc Online Surveys with the consent form attached for each submission. The form collected the following information:

- Consent for testing
- Age
- Sex
- Weight
- Height
- Ethnicity
- Current Medical or Surgical History
- Current Medications
- NLT Data
- Location of Data Collection

All data was anonymously collected, and information was stored with the University of Leeds secure driver, in line with the ethical guidelines and approval.

#### Statistical Analysis

Microsoft Excel was used in the management and formatting of all the data collected. Data analysis was conducted through GraphPad Prism including standard statistical tests and statistical significance calculations.

Statistical analysis was conducted using GraphPad Prism, including standard statistical tests and significance calculations. Microsoft Excel was utilized for data management and formatting. Results are presented as mean ± standard deviation unless otherwise specified.

## Results

### Consort Diagram

### Patient Demographics

Demographic data for the 112 participants (68 female [60.7%]; 44 male [39.3%]; age 35.3 ± 15.1 years) of this cohort are shown in *Table 1*. Most participants were White (69 [61.6%]), with other ethnicities including Asian (9 [8.0%]), Black/Caribbean (4 [3.6%]), Mixed (11 [9.8%]), and Other (19 [17.0%]). Mean BMI was 24.8 ± 4.0 kg/m^2^ (range 17.8-39.4). Thirty-four participants (30.6%) reported at least one medical condition, with asthma (9 [8.0%]), depression (5 [4.5%]), hypertension (4 [3.7%]), and GORD (3 [2.8%]) being most common. Forty-two participants (37.5%) were taking at least one medication, with metformin (5 [4.5%]), COCP (5 [4.5%]), ramipril (4 [3.6%]), salbutamol (4 [3.6%]), and atorvastatin (3 [2.7%]) most frequently reported.

**Table 1:** Table of Demographics

| Characteristic | Value |
| --- | --- |
| Sample Size | 112 |
| **Age (years)** |  |
| Mean ± SD | 35.3 ± 15.1 |
| Median | 31 |
| Range | 18-86 |
| **Sex** |  |
| Female | 68 (60.7%) |
| Male | 44 (39.3%) |
| **Ethnicity** |  |
| White | 69 (61.6%) |
| Asian | 9 (8.0%) |
| Black/Caribbean | 4 (3.6%) |
| Mixed | 11 (9.8%) |
| Other | 19 (17.0%) |
| **BMI (kg/m<sup>2</sup>)** |  |
| Mean ± SD | 24.8 ± 4.0 |
| Median | 24.4 |
| Range | 17.8-39.4 |
| **Medical Conditions** |  |
| Participants with any medical condition | 34 (30.6%) |
| **Most common conditions** |  |
| Asthma | 9 (8.0%) |
| Depression | 5 (4.5%) |
| Hypertension | 4 (3.7%) |
| GORD | 3 (2.8%) |
| **Current Medications** |  |
| Participants on any medication | 42 (37.5%) |
| **Most common medications** |  |
| Metformin | 5 (4.5%) |
| COCP | 5 (4.5%) |
| Ramipril | 4 (3.6%) |
| Salbutamol | 4 (3.6%) |
| Atorvastatin | 3 (2.7%) |

#### NLT HR Data

Heart rate data during the NLT are presented in Table 2. Kolmogorov-Smirnov tests confirmed normal distribution of the HR data. The maximum HR value compared to baseline was used across all 10 minutes of standing for analysis. Mean supine HR for all participants was 71.7 ± 12.8 bpm. Upon assumption of upright posture, the mean HR change was 9.89 ± 8.15 bpm, while the mean sustained HR change (defined as maximum HR maintained over at least 2 consecutive measurements when standing) was 6.33 ± 6.82 bpm.

**Table 2:** Table breaking down NLT test findings. Sustained HR is defined as the maximum HR was maintained over at least 2 consecutive measurements when standing. HR value used is the maximum or minimum value compared to baseline across all 10 minutes of standing. Kolmogorov-Smirnov Tests of Normality were conducted, and data was found to be normally distributed.

| NLT Metrics | Value |
| --- | --- |
| **Lying Down Supine** |  |
| Heart Rate (bpm), Mean ± SD | 71.7 ± 12.8 |
| **Supine to Standing (0-10 min)** |  |
| Average HR change (increase) (bpm), Mean ± SD | 9.89 ± 8.15 |
| Average Sustained HR change (increase) (bpm), Mean ± SD | 6.33 ± 6.82 |

#### NLT BP Data

Blood pressure responses to NLT are shown in *Table 3*. All BP measurements were normally distributed as confirmed by Kolmogorov-Smirnov tests. Both early phase standing (0-3 min) and late phase (4-10 min) standing changes were compared to supine baseline values. Mean supine systolic and diastolic BP were 120.4 ± 12.5 mmHg and 76.3 ± 9.0 mmHg, respectively. During early phase standing, the mean systolic BP increased by 7.55 ± 10.88 mmHg and mean diastolic BP increased by 4.97 ± 8.36 mmHg. In the late phase, the mean systolic BP change decreased by 3.53 ± 10.01 mmHg and diastolic BP change by 3.31 ± 7.64 mmHg.

**Table 3:** Table breaking down NLT Blood Pressure findings. Early Phase are values during the first 3 minutes of standing and Late Phase are values from 4-10 minutes; both changes are compared to lying down baseline value. Kolmogorov-Smirnov Tests of Normality were conducted, and data was found to be normally distributed.

| NLT Metrics | Value |  |
| --- | --- | --- |
| Lying Down Supine |  |  |
| Average Systolic BP (mmHg), Mean ± SD | 120.4 ± 12.5 |  |
| Average Diastolic BP (mmHg), Mean ± SD | 76.3 ± 9.0 |  |
| Supine to Standing | Early Phase (0-3 min) | Late Phase (4-10 min) |
| Average Systolic BP change (increase) (mmHg), Mean ± SD | 7.55 ± 10.88 | 3.53 ± 10.01 |
| Average Diastolic BP change (increase) (mmHg), Mean ± SD | 4.97 ± 8.36 | 3.31 ± 7.64 |

#### NLT Symptoms

Symptoms occurring across all participants during the NLT are presented in *Table 4*. Nine participants (8.0%) reported symptoms upon assumption of upright posture. The most reported symptom was dizziness/unsteadiness (6 [5.4%]), followed by pins and needles/numbness (2 [1.8%]) and back pain (2 [1.8%]).

**Table 4:** Table of symptoms described by participants during the NLT

| Symptoms | Number of Participants (%) |
| --- | --- |
| Participants with any symptoms on standing | 9 (8.0%) |
| Dizziness/unsteadiness | 6 (5.4%) |
| Pins and needles/numbness | 2 (1.8%) |
| Back pain | 2 (1.8%) |

There was n=4 participants who had abnormal NLTs. 1 participant had a systolic BP drop of greater than 20 mmHg, 1 participant had an increase of 30 bpm from lying and 2 participants had persistent tachycardia above 120 bpm. These 4 participants were asymptomatic.

Orthostatic increases in HR during the NLT are shown in *Figure 2*. Mean HR increased immediately upon standing to 7.0 bpm above baseline. This initial HR increase diminished progressively over the 10-minute standing period. A slight plateau was observed between 4-8 min with values ranging from 5.9 bpm to 6.1 bpm above baseline. The HR increase reached its end at 4.8 bpm. The time course demonstrates an initial compensatory HR increase followed by partial recovery but with sustained elevation compared to supine values throughout the test duration.

**Figure 1:**
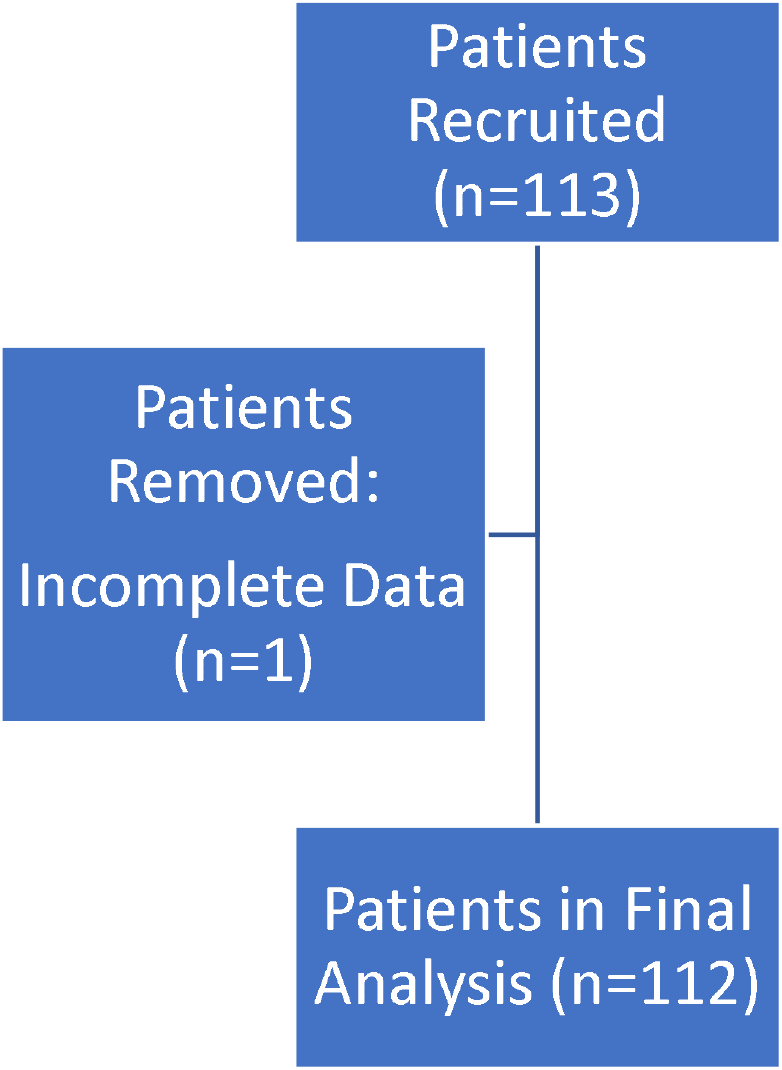
Consort Diagram for patient recruitment. n=1 participants were removed for an incomplete dataset.

**Figure 2:**
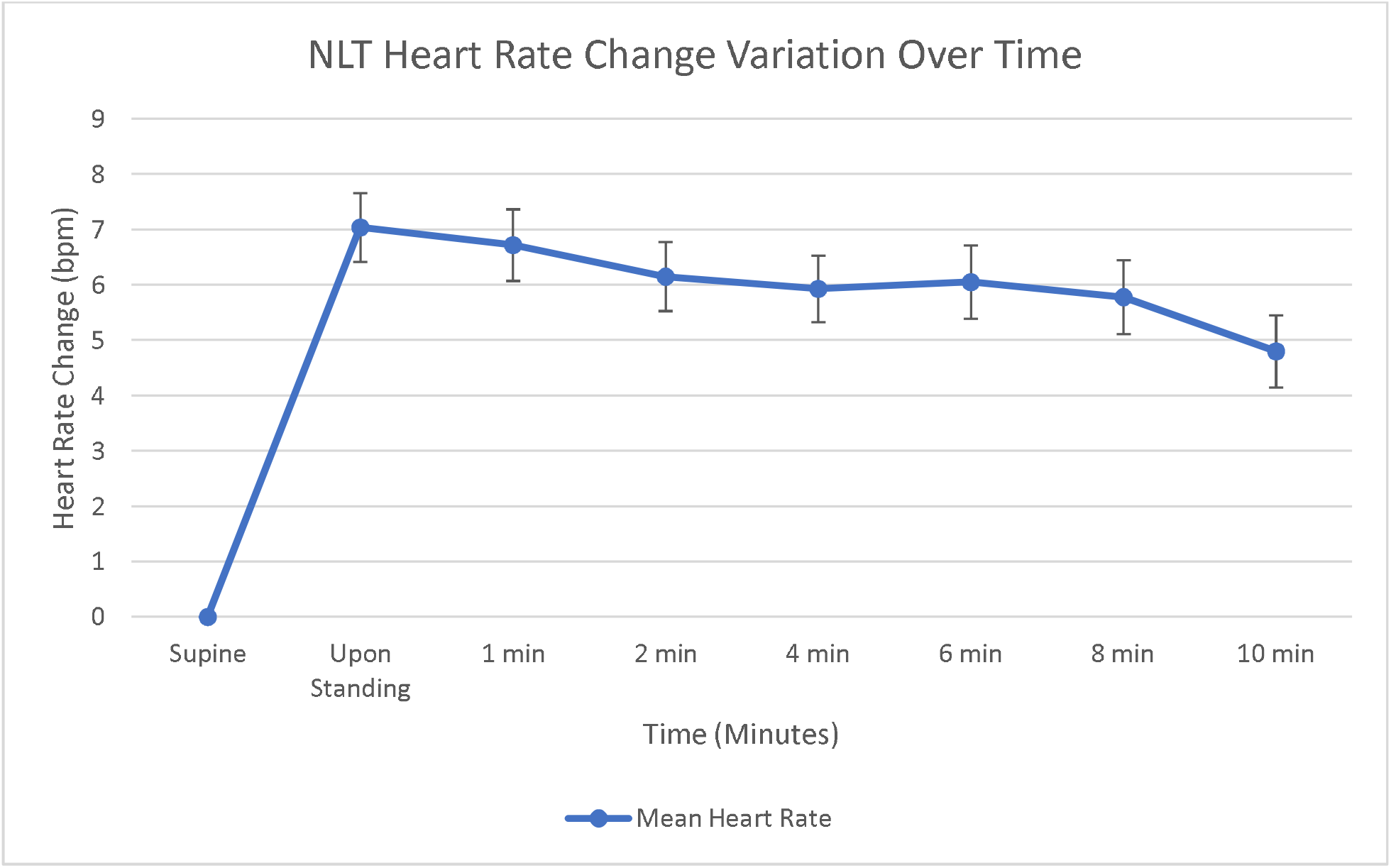
Graph showing mean heart rate change from supine at 1,2,4,6,8 and 10 mins. Standard Error bars.

Blood pressure responses to orthostatic challenge are presented in *Figure 3*. Both systolic and diastolic BP exhibited immediate increases upon standing, with systolic BP showing a greater magnitude of change, 6.7 mmHg, compared to diastolic BP, 4.2 mmHg. Both demonstrated progressive decreases over the 10-minute standing period, with more pronounced declines observed in the early phase (0-3 min) than in the late phase (4-10 min). By test conclusion, systolic BP had decreased to 2.4 mmHg above baseline, while diastolic BP showed similar values at 2.2 mmHg above baseline. There are convergence of systolic BP and diastolic BP increases at approximately 4 minutes. Like HR, BP has an initial increase, followed by a decline and plateau, always remaining above baseline.

**Figure 3:**
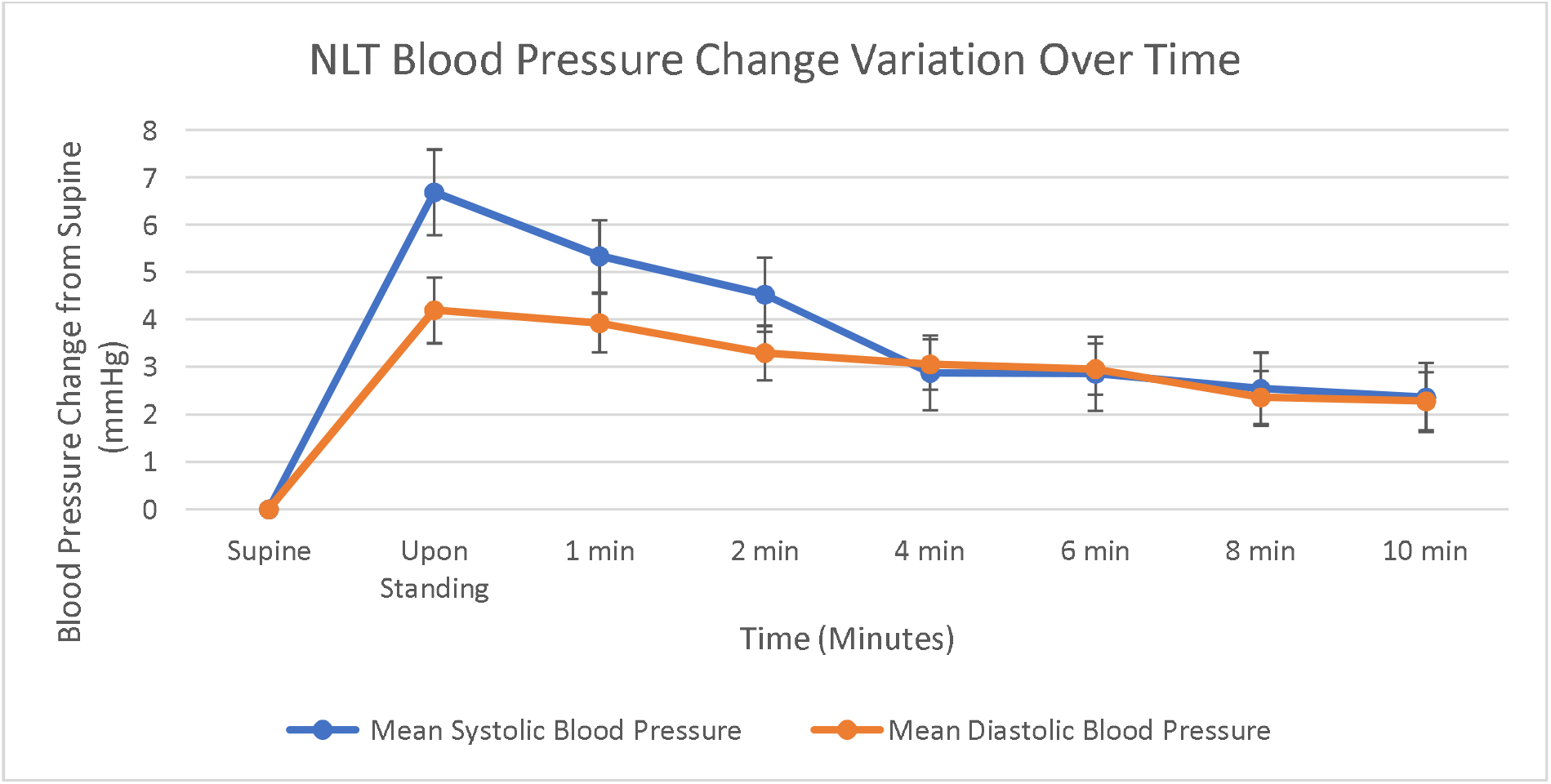
Graph showing mean systolic blood pressure and diastolic blood pressure change from supine at 1,2,4,6,8 and 10 mins. Standard Error bars.

## Discussion

As far as we are aware, this is the first paper within the academic literature to conduct NLTs on a healthy cohort to attain values and understanding of what is to be expected. The assumption from this is that the finding of a negative NLT would help exclude OI in a patient.

Compared with other healthy cohorts for NLTs, the largest group is the healthy population from the study by Lee *et al* in 2024 (36) with a population size of n=50. The gender distribution was similar (64% female in the Lee et al study and 61.6% in this study). The ethnic distribution was similar for the White category in both studies (62%), although differences were evident in the distribution of other ethnicities, particularly in the Mixed/Multiple category (28% in Lee *et al* study compared to 9.8% in this study). The average BMI was nearly identical between the two cohorts (25 ± 5 kg/m^2^ in the Lee et al study and 24.8 ± 4.0 kg/m^2^ in this study) However there was a notable distinction in the age distribution, with Lee *et al’s* population being significantly older on average (48 ± 16 years) compared to this study (35.3 ± 15.1 years). The prevalence of pre-existing conditions also differed (12% in Lee et al study versus 30.6% in this study). Hypertension was present in similar proportions (6% in Lee *et al* study and 3.6% in this study). This study therefore provided a comparative normative data in a younger population that is needed while managing younger individuals with OI.

The normative data from our study can be compared with NLT data collected in the study by Isaac *et al (33)* from 100 LC patients. The mean HR changes in that study was 18.45 ± 9.93 bpm which was significantly different from data collected in this study (unpaired t-test, p<0.0001). This also provides further support for clinical use of NLTs in screening for OI. Once again, the age difference could be a confounding factor in comparison (46.6 years in Isaac et al paper vs 35.6 years in this study) and a direct statistical significance cannot not be calculated.

Our study shows that the NLT is a useful tool to detect OI in LC patients, as it is easy and safe to do either at home or in a clinic. Orthostatic Intolerance and dysautonomia can be missed in those with multiple health conditions and can cause serious problems if left untreated, hence early diagnosis and treatment are important. More research is needed to understand how OI affects LC patients over time. Health professionals who work with LC patients should learn how to assess, interpret, and manage OI and dysautonomia, and have clear guidelines for further care.

This study provides comparative data for using NLT to screen for OI and dysautonomia in health conditions including LC. From this cohort of patients, we have the average HR and BP that one can expect to see in those who do not have dysautonomia or OI. The study provides some reasonable thresholds for HR and BP change beyond which one can expect to see OI symptoms. It can also be argued that the current thresholds for PoTS and OH are based on studies that used HUT test in hospital settings that are being used in NLT and not necessarily validated for NLT. This also explains why some NLT even though convincingly positive for symptoms do not meet the required thresholds for PoTS or OH. Isaac et al in their study explore the alternative threshold options (for example HR change of 20 or 25 in PoTS) in LC (33).

The origins of the NLT goes back to a paper entitled “Cardiovascular deconditioning during space flight and the use of saline as a countermeasure to orthostatic intolerance “(37) whereby the use of a tilt table test and standing test was found to be comparable, which has been further backed up by a paper by Plash *et al;* finding that the diagnosis of PoTS using both techniques was more reliable using a 10-min standing test as the Head Up Tilt (HUT) test had lower specificity with higher false positives in healthy volunteers test (38). This study also suggested that the conventional thresholds used for PoTS diagnosis were based on HUT test and need to lower when using the 10-min stand test. These studies further validate the use of NLT for more widespread use in clinical practice.

The use of NLT can reduce the overall cost in the clinical management for autonomic dysfunction as the NLT is cheap and can be performed in home settings when compared to HUTT hospital test (39). The benefit of the NLT is that it uses only a BP equipment which is readily available to both clinicians and patients and can be conducted in a manner that is easy to understand and replicate across locations with simple instructions. This can aid in management in any health setting including primary care and community healthcare.

When comparing the normative values in a healthy population seen in this study to that of a LC population, we can see that there is significant difference in HR and BP changes that correlates to symptoms during the test. This provides further validation of the presence of dysautonomia in LC and other medical conditions. The test also provides a repeatable objective test that patients and clinicians can relate to and use the test to assess response to interventions and understand condition trajectory (35). This is particularly important when there is currently no single defined biomarker identified for LC which can be used for diagnosis and monitoring.

We chose a younger population in this study to generate normative data that can be used for comparison in LC and other post-infection conditions which are common in this age group. One of the conditions which is related to these conditions and has significant dysautonomia is Myalgic Encephalomyelitis/ Chronic Fatigue Syndrome (ME/CFS). This condition has an early onset (40) and needs normative data to interpret NLT findings. This would allow for a direct comparison and further strengthen any association that may be found with using this NLT for dysautonomia patients and potentially lead to clinical guidelines for LC or any medical condition with dysautonomia syndromes.

One of the limitations of this study is the COVID-19 status of the participants. Even though there is no history of LC or post-infection conditions, it is not known whether had COVID-19 and fully recovered or they truly did not have the infection during the pandemic. This however is not relevant because LC and other post-infection conditions are clinical syndromes and not dependent on having confirmatory lab tests that prove the infection. It is also impractical to find individuals who have not had any infection in the past as almost everyone in the general population is subjected to infections of one type or other and most individuals make a full recovery from infections. The sample we got for this study is as best a normative cohort one can get from general population without any clinical symptoms.

## Author Contributions

MS conceptualised the study and obtained ethical approval. NI and KJ performed the statistical analysis, and NI produced the initial manuscript draft. All authors approved the final manuscript. MS is the corresponding author and guarantor.

## Data Availability

All data produced in the present study are available upon reasonable request to the authors

## References

1. Isaac RO, Corrado J, Sivan M. Detecting Orthostatic Intolerance in Long COVID in a Clinic Setting. International Journal of Environmental Research and Public Health. 2023;20(10):5804.

2. Ahmed H, Patel K, Greenwood DC, Halpin S, Lewthwaite P, Salawu A, et al. Long-term clinical outcomes in survivors of severe acute respiratory syndrome and Middle East respiratory syndrome coronavirus outbreaks after hospitalisation or ICU admission: A systematic review and meta-analysis. J Rehabil Med. 2020;52(5):jrm00063.

3. Khoja O, Silva Passadouro B, Mulvey M, Delis I, Astill S, Tan AL, et al. Clinical Characteristics and Mechanisms of Musculoskeletal Pain in Long COVID. J Pain Res. 2022;15:1729–48.

4. Yilmaz S, Yokusoglu M, Cinar M, Simsek I, Baysan O, Oz BS, et al. Autonomic functions in acrocyanosis assessed by heart rate variability. Eur J Rheumatol. 2014;1(1):18–20.

5. Narasimhan B, Calambur A, Moras E, Wu L, Aronow W. Postural Orthostatic Tachycardia Syndrome in COVID-19: A Contemporary Review of Mechanisms, Clinical Course and Management. Vasc Health Risk Manag. 2023;19:303–16.

6. Blitshteyn S, Whiteson JH, Abramoff B, Azola A, Bartels MN, Bhavaraju-Sanka R, et al. Multi-disciplinary collaborative consensus guidance statement on the assessment and treatment of autonomic dysfunction in patients with post-acute sequelae of SARS-CoV-2 infection (PASC). PM R. 2022;14(10):1270–91.

7. Low PA, Sandroni P, Joyner M, Shen WK. Postural tachycardia syndrome (POTS). Journal of Cardiovascular Electrophysiology. 2009;20(3):352–8.

8. Chandan JS, Brown KR, Simms-Williams N, Bashir NZ, Camaradou J, Heining D, et al. Non-Pharmacological Therapies for Post-Viral Syndromes, Including Long COVID: A Systematic Review. Int J Environ Res Public Health. 2023;20(4).

9. Organisation WH. Post COVID-19 condition (Long COVID) 2023 [cited 2023 18/05/2023]. Available from: https://www.who.int/europe/news-room/fact-sheets/item/post-covid-19-condition.

10. Greenhalgh T, Sivan M, Perlowski A, Nikolich J. Long COVID: a clinical update. Lancet. 2024;404(10453):707–24.

11. Office of National Statistics. Prevalence of ongoing symptoms following coronavirus (COVID-19) infection in the UK: 1 June 2022. London: ONS; 2022.

12. Greenhalgh T, Sivan M, Delaney B, Evans R, Milne R. Long covid-an update for primary care. BMJ. 2022;378:e072117.

13. Corrado J, Iftekhar N, Halpin S, Li M, Tarrant R, Grimaldi J, et al. HEART Rate Variability Biofeedback for LOng COVID Dysautonomia (HEARTLOC): Results of a Feasibility Study. Advances in Rehabilitation Science and Practice. 2024;13:27536351241227261.

14. Acanfora D, Nolano M, Acanfora C, Colella C, Provitera V, Caporaso G, et al. Impaired Vagal Activity in Long-COVID-19 Patients. Viruses. 2022;14(5):13.

15. Iftekhar N, Sivan M. Venous insufficiency and acrocyanosis in long COVID: dysautonomia. Lancet. 2023;402(10401):e9.

16. Yong SJ. Long COVID or post-COVID-19 syndrome: putative pathophysiology, risk factors, and treatments. Infectious Diseases. 2021;53(10):737–54.

17. Dani M, Dirksen A, Taraborrelli P, Torocastro M, Panagopoulos D, Sutton R, et al. Autonomic dysfunction in ‘long COVID’: rationale, physiology and management strategies. Clin Med (Lond). 2021;21(1):e63–e7.

18. Low PA, Opfer-Gehrking TL, Textor SC, Schondorf R, Suarez GA, Fealey RD, et al. Comparison of the postural tachycardia syndrome (POTS) with orthostatic hypotension due to autonomic failure. J Auton Nerv Syst. 1994;50(2):181–8.

19. Ashton RE, Philips BE, Faghy M. The acute and chronic implications of the COVID-19 virus on the cardiovascular system in adults: A systematic review. Prog Cardiovasc Dis. 2023;76:31–7.

20. Wallukat G, Hohberger B, Wenzel K, Fürst J, Schulze-Rothe S, Wallukat A, et al. Functional autoantibodies against G-protein coupled receptors in patients with persistent Long-COVID-19 symptoms. J Transl Autoimmun. 2021;4:100100.

21. Seeley MC, Gallagher C, Ong E, Langdon A, Chieng J, Bailey D, et al. High Incidence of Autonomic Dysfunction and Postural Orthostatic Tachycardia Syndrome in Patients with Long COVID: Implications for Management and Health Care Planning. Am J Med. 2025;138(2):354–61.e1.

22. Kavi L. Postural tachycardia syndrome and long COVID: an update. British Journal of General Practice. 2022;72(714):8–9.

23. Harris CI. COVID-19 Increases the Prevalence of Postural Orthostatic Tachycardia Syndrome: What Nutrition and Dietetics Practitioners Need to Know. J Acad Nutr Diet. 2022;122(9):1600–5.

24. Eastin E, Machnik J, Larsen N, Stiles L, Seliger J, Geng L, et al. Evaluating Long-Term Autonomic Dysfunction and Functional Impacts of Long COVID: A Follow-Up Study (P1-7.004). Neurology. 2025;104(7_Supplement_1):4290.

25. Kato A, Tokumasu K, Yamamoto K, Otsuka Y, Nakano Y, Honda H, et al. Clinical and endocrine features of orthostatic intolerance detected in patients with long COVID. Scientific Reports. 2024;14(1):17025.

26. Hira R, Baker JR, Siddiqui T, Ranada SI, Soroush A, Karalasingham K, et al. Objective Hemodynamic Cardiovascular Autonomic Abnormalities in Post-Acute Sequelae of COVID-19. Canadian Journal of Cardiology. 2023;39(6):767–75.

27. Carod-Artal FJ. Infectious diseases causing autonomic dysfunction. Clin Auton Res. 2018;28(1):67–81.

28. Erdal Y, Atalar AC, Gunes T, Okluoglu T, Yavuz N, Emre U. Autonomic dysfunction in patients with COVID-19. Acta Neurologica Belgica. 2022;122(4):885–91.

29. Goldstein DS, Robertson D, Esler M, Straus SE, Eisenhofer G. Dysautonomias: clinical disorders of the autonomic nervous system. Ann Intern Med. 2002;137(9):753–63.

30. Oakley I, Emond L. Diabetic cardiac autonomic neuropathy and anesthetic management: review of the literature. Aana j. 2011;79(6):473–9.

31. Lee J, Vernon SD, Jeys P, Ali W, Campos A, Unutmaz D, et al. Hemodynamics during the 10-minute NASA Lean Test: evidence of circulatory decompensation in a subset of ME/CFS patients. J Transl Med. 2020;18(1):314.

32. Lee C, Greenwood DC, Master H, Balasundaram K, Williams P, Scott JT, et al. Prevalence of orthostatic intolerance in long covid clinic patients and healthy volunteers: A multicenter study. J Med Virol. 2024;96(3):e29486.

33. Isaac RO, Corrado J, Sivan M. Detecting Orthostatic Intolerance in Long COVID in a Clinic Setting. Int J Environ Res Public Health. 2023;20(10).

34. Sivan M, Greenhalgh T, Darbyshire JL, Mir G, O’Connor RJ, Dawes H, et al. LOng COvid Multidisciplinary consortium Optimising Treatments and servIces acrOss the NHS (LOCOMOTION): protocol for a mixed-methods study in the UK. BMJ Open. 2022;12(5):e063505.

35. Sivan M CJ, Mathias C. The adapted Autonomic Profile (aAP) home-based test for the evaluation of neuro-cardiovascular autonomic dysfunction. Adv Clin Neurosci Rehabil 2022. 2022.

36. Lee C, Greenwood DC, Master H, Balasundaram K, Williams P, Scott JT, et al. Prevalence of orthostatic intolerance in long covid clinic patients and healthy volunteers: A multicenter study. Journal of Medical Virology. 2024;96(3):e29486.

37. Bungo MW, Charles JB, Johnson PC, Jr. Cardiovascular deconditioning during space flight and the use of saline as a countermeasure to orthostatic intolerance. Aviat Space Environ Med. 1985;56(10):985–90.

38. Plash WB, Diedrich A, Biaggioni I, Garland EM, Paranjape SY, Black BK, et al. Diagnosing postural tachycardia syndrome: comparison of tilt testing compared with standing haemodynamics. Clinical science (London, England : 1979). 2013;124(2):109–14.

39. Asahina M, Akaogi Y, Yamanaka Y, Koyama Y, Hattori T. Differences in skin sympathetic involvements between two chronic autonomic disorders: multiple system atrophy and pure autonomic failure. Parkinsonism Relat Disord. 2009;15(5):347–50.

40. Bae J, Lin JS. Healthcare Utilization in Myalgic Encephalomyelitis/Chronic Fatigue Syndrome (ME/CFS): Analysis of US Ambulatory Healthcare Data, 2000–2009. Front Pediatr. 2019;7:185.

